# A Multimodal Multiomics Machine Learning (MMM) approach for biomarker discovery and acceleration of clinical trial readiness for childhood-onset neurological disorders

**DOI:** 10.64898/2026.07.21.26358463

**Authors:** Audrey K.S. Soo, Jenny Hällqvist, Kiran Seunarine, Robert Spaull, Ivan Doykov, Sara Guttmann, Kathleen Gorman, Apostolos Papandreou, Tian Luo, Yi Wang, Maya Thomas, Sangeetha Yoganathan, Evangeline Wassmer, Belén Pérez-Dueñas, Alejandra Darling, Nardo Nadocci, Giovanna Zorzi, Boriana Büchner, Thomas Klopstock, Amitav Parida, Francesca Magrinelli, Kailash P. Bhatia, Allison Gregory, Katrina Wakeman, Penelope Hogarth, Susan Hayflick, Amanda Heslegrave, Henrik Zetterberg, Wendy Heywood, Asthik Biswas, Ulrike Löbel, Kshitij Mankad, Jan Sedlacik, Sniya Sudhakar, Christopher Clark, Kevin Mills, Manju A. Kurian, PLAN research consortium

**Affiliations:** Developmental Neurosciences, Zayed Centre for Research into Rare Disease in Children, Great Ormond Street–Institute of Child Health (GOS-ICH), University College London (UCL), London, England; Department of Neurology, Great Ormond Street Hospital for Children (GOSH), London, England; University College London Great Ormond Street Hospital Institute of Child Health, London, UK; Children’s Hospital of Fudan University, Shanghai, China; Christian Medical College, Vellore, Tamil Nadu, India; Birmingham Children’s Hospital, Birmingham, England; Vall d’Hebron University Hospital and Biomedical Research Networking Centre on Rare Diseases (CIBERER), Barcelona, Spain; Neurology Department, Metabolic Unit and Movement Disorders Unit, Sant Joan de Déu Barcelona Children’s Hospital, Barcelona, Spain; Fondazione I.R.C.C.S. Instituto Neurologico Carlo Besta, Milan, Italy; Department of Neurology, Friedrich-Baur-Institute, LMU Hospital, Ludwig-Maximilians-Universität (LMU) München, Munich, Germany; German Center for Neurodegenerative Diseases (DZNE) and Munich Cluster for Systems Neurology (SyNergy), Munich, Germany; Department of Clinical and Movement Neurosciences, UCL Queen Square Institute of Neurology, London, England; Oregon Health and Science University, Oregon, USA; UCL UK Dementia Research Institute, London, England; UCL Queen Square Institute of Neurology, London, England; Department of Psychiatry and Neurochemistry, Institute of Neuroscience and Physiology, the Sahlgrenska Academy at the University of Gothenburg, Mölndal, Sweden; Clinical Neurochemistry Laboratory, Sahlgrenska University Hospital, Mölndal, Sweden; Hong Kong Center for Neurodegenerative Diseases, Clear Water Bay, Hong Kong, China; Wisconsin Alzheimer’s Disease Research Center, University of Wisconsin School of Medicine and Public Health, University of Wisconsin-Madison, Madison, WI 53792, USA; Department of Neuroradiology, GOSH, London, England; Robert Steiner MR Facility, Medical Research Council Laboratory of Medical Sciences, Hammersmith Hospital Campus and Mansfield Centre for Innovation, Imaging Sciences, Institute of Clinical Sciences, Imperial College London, Hammersmith Hospital Campus, London, UK

## Abstract

**Background:** Childhood neurodegenerative disorders are usually rare, genetic, and life-limiting. Whilst targeted approaches present huge potential, significant hurdles include disease rarity, geographical dispersion of patients, funding, clinical trial design, and execution. Crucially, the paucity of robust biomarkers and objective measures of disease progression hampers evaluation of efficacy, drug development and regulatory approval. To address this paradigm, we developed a Multimodal Multiomics Machine Learning (MMM) framework, integrating large-scale, multi-source patient datasets to generate quantitative metrics for disease stratification and longitudinal tracking. We applied MMM to *PLA2G6*-associated neurodegeneration (PLAN), an ultra-rare condition currently lacking validated biomarkers, where precision gene therapy approaches are at an advanced preclinical stage.

**Methods:** A large, single time-point international natural history study (n = 310) was conducted alongside development of a disease-specific rating scale (CoPLAN-DRS), prospective longitudinal neuroimaging, and multiomic biomarker discovery. Machine learning methods were applied to the integrated dataset.

**Results:** Kaplan-Meier analyses enabled estimates for survival and time to loss of ambulation. Multiple clinical, radiological, and biofluid biomarkers were identified, clearly correlating with disease progression. The CoPLAN-DRS and brain MRI Quantitative Susceptibility Mapping showed strong positive correlation with age (rho = 0.69, 0.96 respectively). Nicastrin, a critical structural component of the gamma-secretase complex in Amyloid Precursor Protein (APP) processing, was identified as a novel biomarker. Neurofilament light levels showed strong negative correlation with disease progression (rho = −0.74). The complex multi-dimensional dataset was distilled into a simplified, clinically intuitive Digital Disease Dashboard (DDD), enabling real-time visualisation of disease severity.

**Conclusions:** Our study highlights the clinical utility of MMM in integrating multi-dimensional data from rare disease cohorts, delivering an unbiased, data-driven, optimised biomarker set. Condensing this into the DDD provides a pragmatically useful tool for clinicians, facilitating longitudinal tracking of disease. The MMM and DDD have accelerated clinical-trial readiness for PLAN, and potentially applicable to a broad range of neurogenetic disorders.

**RESEARCH IN CONTEXT:** *Evidence before this study:* *PLA2G6*-associated neurodegeneration (PLAN) is a devastating rare, genetic neurodegenerative disorder. Young children affected by PLAN often appear to be developing normally initially before rapid motor and cognitive deterioration, losing any previously gained skills like walking, speaking, and feeding independently. This is a life-limiting disorder without any effective treatments at present, although gene therapy is in advanced stages of preclinical development. The lack of in-depth understanding of the disease spectrum and paucity of biomarkers could potentially hamper the clinical translation of gene therapy and other novel precision therapies.

*Added value of this study:* This study describes a unique approach, harnessing technological advances and machine learning methodology to optimise big data generated from patients, even for relatively small datasets from rare disease cohorts. As proof-of-concept, a Multimodal Multiomics Machine Learning (MMM) strategy was applied to a patient cohort affected by PLAN, for which there are currently no established reliable biomarkers. A machine learning approach was used to distil the complex raw datasets into an accessible platform for applying to clinical practice. The MMM approach has: - identified the best discriminatory biomarkers that differentiate PLAN patients from controls thereby aiding diagnosis
- identified a range of robust biomarkers that reflect disease severity and progression, aiding longitudinal monitoring and prognostication
- objectively assembled an array of the best data-driven optimised biomarkers as potential PLAN outcome measures for upcoming clinical trials We have also developed the Digital Disease Dashboard (DDD), a new visual tool for clinical application that condenses large-scale multidimensional datasets into a single intuitive platform and estimates disease severity at a specific timepoint. The DDD therefore bridges the gap between complex bioinformatics and real-world clinical decision making, offering a powerful new way of communicating disease severity and trajectory, tracking individual patients longitudinally over time.

*Implications of all the available evidence:* This MMM toolkit is broadly applicable, not only to rare, neurological disorders like PLAN but also to other childhood-onset genetic disorders. This data-driven, translational approach to generate and utilise big data will rationalise biomarker discovery, providing better measures of disease progression, thereby accelerating clinical trial readiness and development of more effective therapies.

Graphical abstract

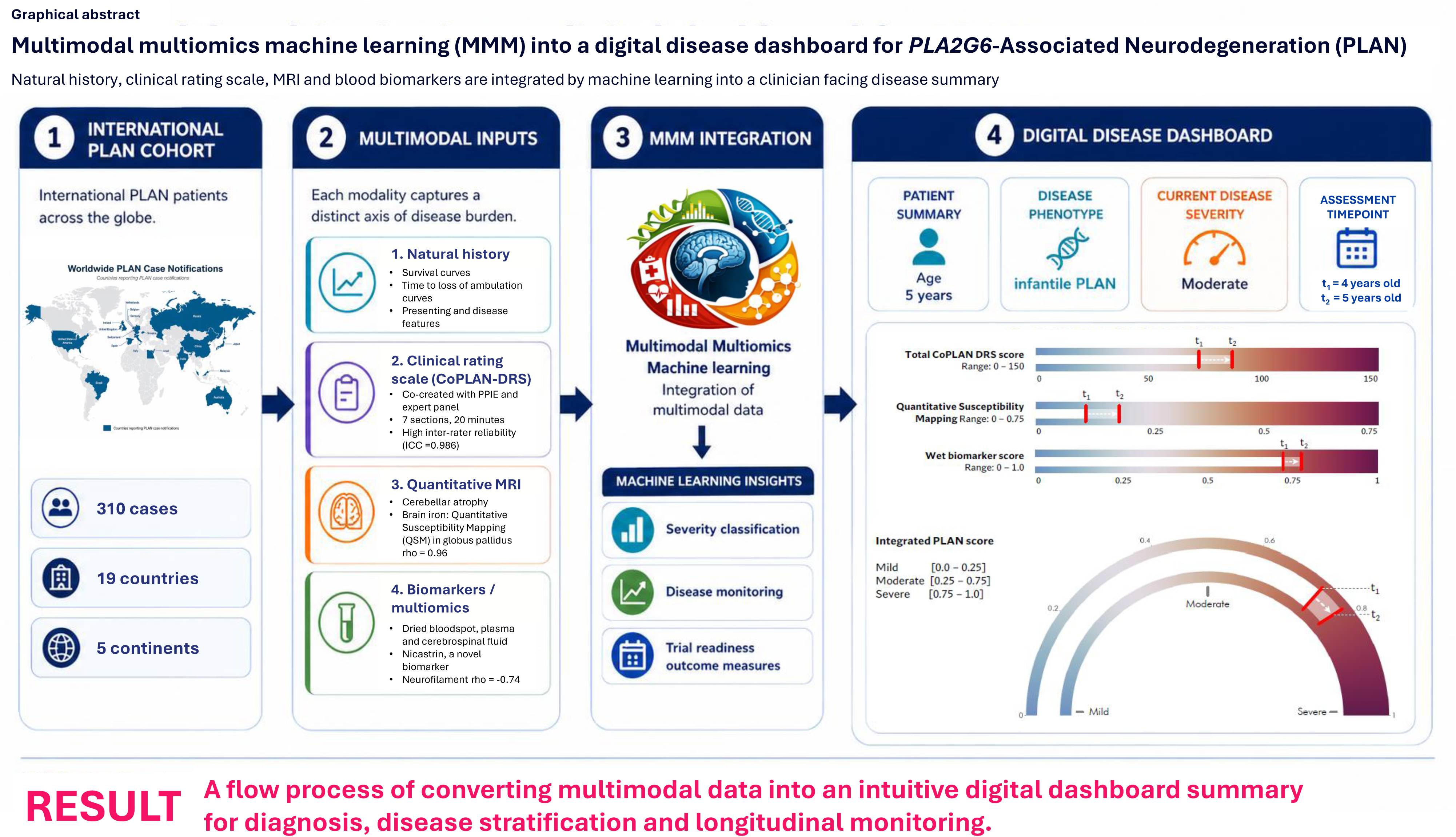

## INTRODUCTION

The vast majority of rare diseases are genetic, life-limiting, and without disease-modifying treatments. They often begin in childhood and involve the nervous system. Whilst targeted genetic and precision therapy approaches hold huge potential for these monogenic disorders, significant hurdles exist, related to disease rarity. Limited numbers of geographically-dispersed patients makes it difficult to undertake comprehensive natural history studies and ensure adequate statistical powering for clinical trials, that also account for phenotypic pleiotropy.^1,2^ Crucially, the lack of objective, robust biomarkers to serve as clinically-relevant, reliable, and reproducible outcome measures for clinical trials hampers the evaluation of efficacy, long-term drug development, and regulatory approval.^3,4^

One condition that epitomises the challenges faced by rare diseases is *PLA2G6*-associated neurodegeneration (PLAN), an ultra-rare form of Neurodegeneration with Brain Iron Accumulation (NBIA) caused by biallelic mutations in *PLA2G6*.^5, 6^ The PLAN continuum comprises late-infantile, juvenile, and adult-onset forms of disease (iPLAN, jPLAN, and aPLAN respectively), though there are currently no consensus definitions for these subgroups.^7^ Other classification terms in use include Infantile Neuroaxonal Dystrophy (INAD), atypical Neuroxonal Axonal Dystrophy (aNAD), *PLA2G6*–related dystonia-parkinsonism, and PARK14.^7^ Affected patients develop motor and cognitive regression and progressive neurodegeneration, with widespread accumulation of tau, Lewy bodies, and neuroaxonal spheroids^8^, ultimately leading to premature death. The commonest form of PLAN (iPLAN) manifests in the first few years of life and is rapidly progressive, often with death before adolescence.

Like PLAN, the majority of childhood-onset neuroregressive disorders are rare, genetic, life-limiting, and without disease-modifying treatments. The lack of robust biomarkers makes it challenging to objectively quantify disease progression and to measure the efficacy of any potential treatment. The development of such biomarkers has been identified as a research priority for childhood developmental regression.^9^

To address these challenges, we have developed a unique Multimodal Multiomics Machine learning (MMM) strategy. The MMM approach harnesses technological advances and machine learning methodology to optimise big data generated from patients, even for relatively small datasets from rare disease cohorts. By generating integrated multi-source patient datasets, MMM aims to improve disease stratification, identify robust disease biomarkers, develop clinical outcome measures, and accelerate clinical trial readiness. The complex raw datasets were distilled into an easily interpretable Digital Disease Dashboard (DDD) to provide an accessible and interpretable platform for clinical decision-making and research. PLAN was chosen as a prototype for this approach, as a viral vector-based gene therapy strategy is in the late stages of preclinical development^10,11^ and there are currently no disease-tailored biomarkers or reliable clinical outcome measures to use in upcoming clinical trials. PLAN provides a particularly valuable model for assessing MMM, given its overlap with prevalent neurodegenerative diseases like Alzheimer’s and Parkinson’s disease. We believe that this pipeline represents a key step towards next-generation disease stratification, with tools capable of monitoring disease progression and therapeutic response in real time, accelerating precision therapy for patients with rare diseases.

## METHODS

The study was approved by the host institution’s review board and national research ethics committees (References: 15NM14, 20/LO/0715, 13/LO/0168). Written informed consent was obtained from study participants and/or their parents, guardians, or consultees.

A large international, single time-point natural history study was conducted using a standardised proforma to collect clinical information on PLAN patients. Age at onset of neuroregression was used to classify PLAN subtype: iPLAN < 3 years; jPLAN 3-18 years; aPLAN >18 years. Patients were invited to participate in development of the disease-specific clinical rating scale, prospective quantitative neuroimaging and biomarker studies.

Childhood-onset PLAN Disease Rating Scale (CoPLAN-DRS) was co-created with a panel of 11 international experts (Delphi consensus) and 10 PLAN families. A single assessor (A.K.S.S.) performed standardised CoPLAN-DRS assessments on patients from 3 international centres. Consented video recordings of the assessment were reviewed by two other paediatric neurologists for independent scoring.

A prospective, single-site, longitudinal brain magnetic resonance imaging (MRI) research study was performed; all patients were scanned on a 3-Tesla Siemens Magnetom Prisma scanner using sequences that included 3D T1-weighted images for volumetrics and T2*-weighted images to compute Quantitative Susceptibility Mapping (QSM) values, a surrogate marker of brain iron accumulation.^12^ Anonymised control scans obtained previously on the same MRI scanner from developmentally normal children with no known neurological disorder were utilised for comparison.

Longitudinal dried blood spots (DBS) and blood plasma samples were collected from PLAN patients. Control samples were obtained from developmentally normal children undergoing routine minor elective surgery. Cerebrospinal fluid (CSF) samples from PLAN patients and controls, previously stored for clinical or research purposes, were also utilised. Proteomic and lipidomic analysis was conducted using mass spectrometry (MS). Single molecular array (Simoa^®^) and proximity extension assay technology using Olink^®^ Target 96 Neuro Exploratory and Neurology panels were also undertaken.

Machine learning methods were applied to the dataset. Principal component analysis (PCA) and orthogonal projection to latent structures discriminant analysis (OPLS-DA) was applied to the combined multiomics, multimodal dataset. A logistic regression model was constructed to discriminate between iPLAN and control samples.

## RESULTS

### PLAN Natural History Study (PLAN-NHiS)

In total, 310 PLAN cases from 19 countries were included (**Figure 1A**). The largest phenotypic group was iPLAN (232, 75%) then jPLAN (62, 20%) and aPLAN (16, 5%) (**Table 1**). There were significant differences in Kaplan-Meier survival curves for the three phenotypic groups (p<0.001) (**Figure 1B**). Median age of survival for iPLAN was 11 years, jPLAN was 39 years and all aPLAN patients were alive at the point of data collection.

**Figure 1:**
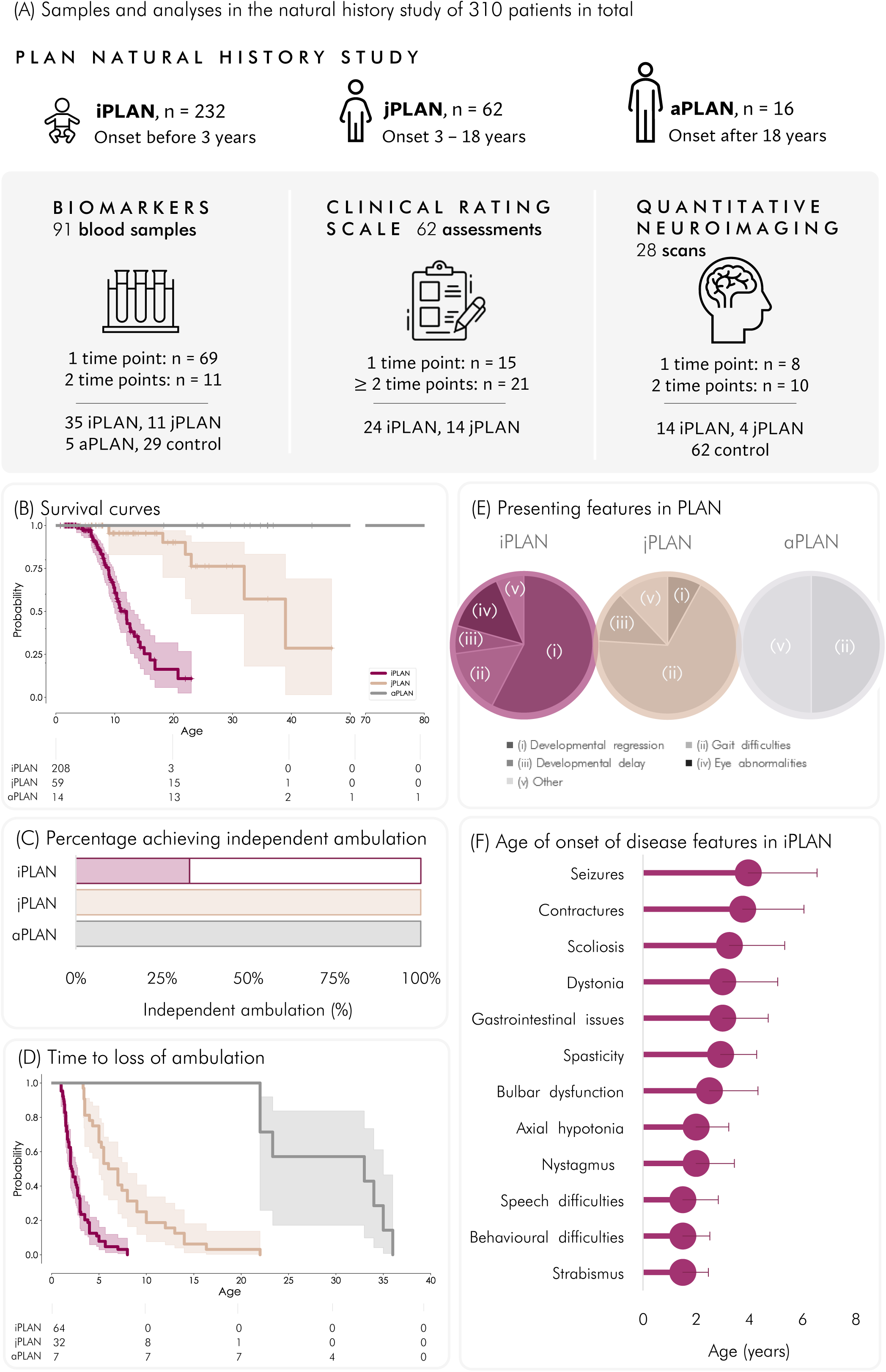
PLAN Natural History Study (PLAN-NHiS) results. **(A)** Summary of study. PLAN patients who entered the single-time point PLAN-NHiS were invited to participate in the prospective biomarker, clinical rating scale and quantitative neuroimaging streams of the study, at either single or multiple timepoints. **(B)** Kaplan Meier survival curves according to PLAN subgroup. The three phenotypic groups were compared using logrank test for trend and were statistically significantly different from each other (p<0.0001). **(C)** Percentage of PLAN patients in each phenotypic group who achieved independent ambulation. Only 1/3 of iPLAN patients ever achieve independent ambulation whereas all jPLAN and aPLAN patients achieved this motor milestone. **(D)** Kaplan Meier plot showing time to loss of ambulation according to PLAN phenotype. The curves for the three phenotypic groups were compared using logrank test for trend and were statistically significantly different from each other (p<0.0001). **(E)** Main presenting features for PLAN according to phenotype. The most common presenting feature for iPLAN was developmental regression, whereas for jPLAN it was gait difficulties and for aPLAN it was either gait difficulties or other features (mainly psychiatric). **(F)** Box and whisker graph showing the age of onset of different disease features in the iPLAN group. Whiskers constructed using the Tukey method.

**Table 1:** Study participants.

|  | PLAN | Controls |
| --- | --- | --- |
| **Natural history study** |  |  |
| Phenotypic PLAN groups |  |  |
| Total | 310 (100) |  |
| Infantile PLAN onset < 3 years (%) | 232 (75) |  |
| Juvenile PLAN onset 3 – 18 years (%) | 62 (20) |  |
| Adult PLAN onset > 18 years (%) | 16 (5) |  |
| Country of notification |  |  |
| High-income (%) | 206 (66) |  |
| Upper-middle income (%) | 62 (20) |  |
| Lower middle income (%) | 42 (14) |  |
| Sex |  |  |
| Female (%) | 153 (49) |  |
| Male (%) | 151 (49) |  |
| Unknown (%) | 6 (2) |  |
| Consanguinity |  |  |
| No (%) | 191 (62) |  |
| Yes (%) | 104 (34) |  |
| Unknown (%) | 13 (4) |  |
| PLA2G6 genetic mutation type |  |  |
| Homozygous mutations (%) | 147 (47) |  |
| Compound heterozygous mutations (%) | 140 (45) |  |
| Other (1 variant, 3 or more variants or unknown) (%) | 15 (7) |  |
| **Clinical rating scale** |  |  |
| Age range |  |  |
| iPLAN (median) | 1.0 – 13.7 (5.4) |  |
| jPLAN (median) | 7.6 – 22.8 (9.0) |  |
| Number of participants |  |  |
| iPLAN total | 24 |  |
| 1 assessment only | 9 |  |
| 2 or more assessments | 15 |  |
| jPLAN total | 14 |  |
| 1 assessment only | 9 |  |
| 2 or more assessments | 5 |  |
| **Neuroimaging** |  |  |
| Number of scans utilised for volumetric analysis | 30 | 62 |
| Number of scans utilised for QSM analysis | 28 | 17 |
| **Biomarker samples** |  |  |
| Dried blood spots |  |  |
| Discovery cohort: untargeted proteomics | 14 | 6 |
| Validation cohort: targeted proteomics | 58 | 13 |
| Targeted lipidomics | 55 | 13 |
| Plasma |  |  |
| Simoa<sup>®</sup> analysis | 47 | 13 |
| Olink<sup>®</sup> analysis | 43 | 13 |
| Cerebrospinal fluid |  |  |
| Simoa<sup>®</sup> testing | 9 | 11 |
| Urine | 28 | 11 |

Only 1/3 (34%) of iPLAN patients achieved independent ambulation whereas all jPLAN and aPLAN patients achieved this developmental milestone (**Figure 1C**). The iPLAN group lost independent ambulation significantly earlier than jPLAN and aPLAN groups (median age iPLAN 2.1 years, jPLAN 6.5 years, aPLAN 28.2 years, p<0.001) (**Figure 1D**).

Features evident on first clinical presentation were reported for 250 cases. Median age at symptom onset differed significantly by subtypes (iPLAN 1.3years, jPLAN 3.0years, aPLAN 20.5years) (p<0·001). The commonest presenting feature for iPLAN was developmental regression (106/184, 58%) whereas gait difficulties and frequent falls were more commonly reported for jPLAN (34/50, 68%) and aPLAN (8/16, 50%) (**Figure 1E**). Characteristic disease features evolved in iPLAN over time (**Figure 1F**).

### CoPLAN-DRS

The newly developed CoPLAN-DRS has 7 sections and maximum score of 150, with higher scores reflecting greater disease severity. 62 CoPLAN-DRS assessments were completed on 38 PLAN patients; 20 patients had 2 longitudinal assessments at least 12 months apart (**Table 1**). The annualised CoPLAN-DRS score in iPLAN patients calculated using a mixed effects model was estimated to be 7.2 points per year. The intraclass correlation coefficient (ICC) between the 3 raters was 0.98 (95% CI 0.97-0.99, p<0.001). There was a significant positive correlation between CoPLAN-DRS score and age in iPLAN (rho = 0.69, p<0.01) and jPLAN (rho = 0.60, p<0.01) (**Figure 2A**).

**Figure 2:**
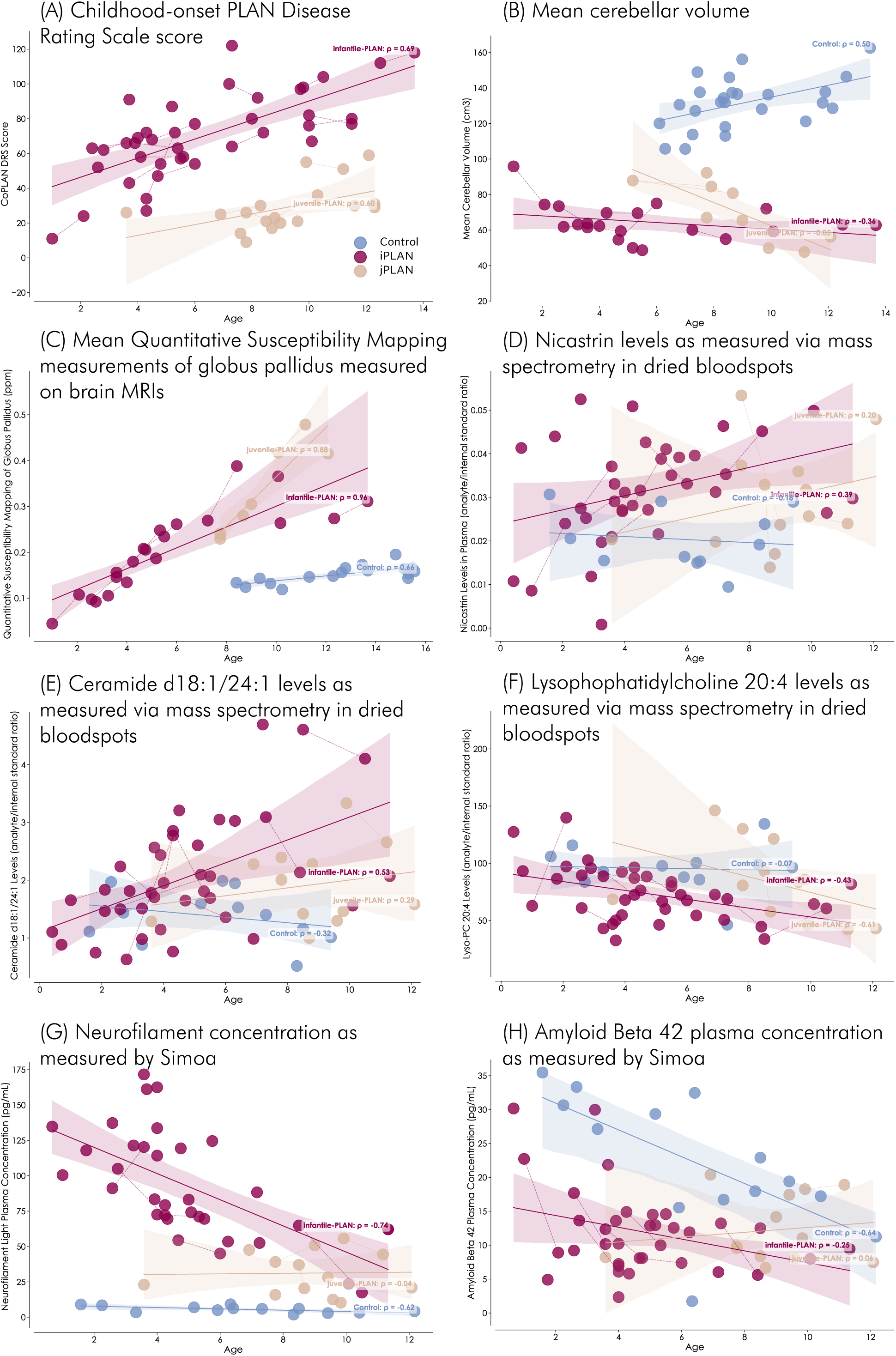
Multimodal multiomic metrics that correlate with age. **(A)** Childhood-onset PLAN Disease Rating Scale (CoPLAN-DRS) score **(B)** Mean cerebellar volume (cm^3^) **(C)** Mean Quantitative Susceptibility Mapping (QSM) measurements of globus pallidus measured on brain MRIs (parts per million, ppm) **(D)** Nicastrin levels as measured via mass spectrometry on dried blood spots **(E)** Ceramide d18:1/24:1 levels as measured via mass spectrometry on dried blood spots **(F)** Lysophophatidylcholine 20:4 levels as measured via mass spectrometry on dried blood spots **(G)** Neurofilament concentration (pg/ml) as measured with Simoa^®^ **(H)** Amyloid Beta 40 plasma concentration (pg/ml) as measured with Simoa^®^ Age acts as surrogate for disease progression given the progressive, life-limiting, neurodegenerative nature of iPLAN and jPLAN. Each dot represents a single measurement. Thin lines connecting 2 or more dots indicate repeated longitudinal measures in individual patients. The thick lines represented linear regression. The shaded band around the regression line represents the 95% confidence interval. Each phenotype is annotated with the Spearman correlation coefficient rho. Maroon = infantile-PLAN, brown = juvenile-PLAN, blue = controls.

### Quantitative Brain Magnetic Resonance Imaging (MRI)

28 prospective brain MRI scans were performed in 18 PLAN patients; 10 patients had a baseline scan followed by an interval scan 12-16 months later. The volumes of the total brain, total white matter, total grey matter, posterior corpus callosum, and cerebellum were significantly reduced in PLAN patients compared to age-matched controls (**Figures 2B**). Brain iron levels, as measured by QSM, of PLAN patients significantly positively correlated with age in the globus pallidus (GP) (rho = 0.93, p<0.001) (**Figure 2C**) and substantia nigra (SN) (rho = 0.84, p<0.001). PLAN QSM values were significantly higher than controls and increased over time in all individuals who had interval scans.

### Lipids and Proteins

Various patient and control biofluid samples were used for biomarker discovery (**Table 1**). Discovery proteomics performed on 20 DBS samples (14 PLAN, 6 controls), identified 52 proteins that were significantly upregulated or downregulated in PLAN, compared to controls. Nicastrin was the most significantly upregulated protein. For validation, these potential biomarkers were translated into a targeted and multiplexed proteomic test and used to analyse 71 additional DBS samples (58 PLAN, 13 controls). This assay confirmed disease-specific upregulation of nicastrin (p<0.001) (**Figure 2D**).

The greatest difference in lipid levels between controls and iPLAN were dihexosylceramide (Hex2Cer) and lysophosphatidylcholine (LPC), especially C24 species (**Figures 2E and 2F**). Analysis of CoPLAN-DRS score and lipids found that the strongest positive correlation was with nervonic (C24:1) acid ceramide. The difference between the control and iPLAN groups was more strongly pronounced when evaluating the ratios between lyso- and diacylated lipids (LPC C20:4/PC C36:4 ratio, p = 7.8E^-7^). When modelling LPC as a function of its diacylated precursor phosphatidylcholine (PC), there was a significantly different linear relationships between the control and iPLAN groups (p = 0.0001).

Simoa^®^-based analysis revealed that all seven biomarkers tested in plasma and 4/5 biomarkers tested in CSF were significantly different in PLAN patients compared to age-matched controls (**Figures 2G and 2H)**. There was a disease-specific significant negative correlation between plasma neurofilament light (NfL) levels and age (rho = −0.74, p<0.001 **Figure 2G**). Analysis by Olink^®^ showed that NfL was also significantly raised in iPLAN and jPLAN compared to controls.

### Machine Learning

To evaluate associations between CoPLAN-DRS score, biomarkers, natural history data, and MRI metrics, an OPLS model using iPLAN and jPLAN samples was constructed, with CoPLAN-DRS score as the response. The model was significant (P = 4.2 E), identifying IL12, lysophosphatidylcholine, and cerebellar volume as principal contributors. An OPLS-DA classifier trained on baseline iPLAN and control samples (CV ANOVA P = 2.9E; permutation p<0.001, **Figure 3A**) accurately classified all follow-up iPLAN samples. 9/14 jPLAN samples were predicted as iPLAN, 5/14 remained unclassified. The first orthogonal component captured age-related variance while the second grouped follow-up samples by individual.

**Figure 3:**
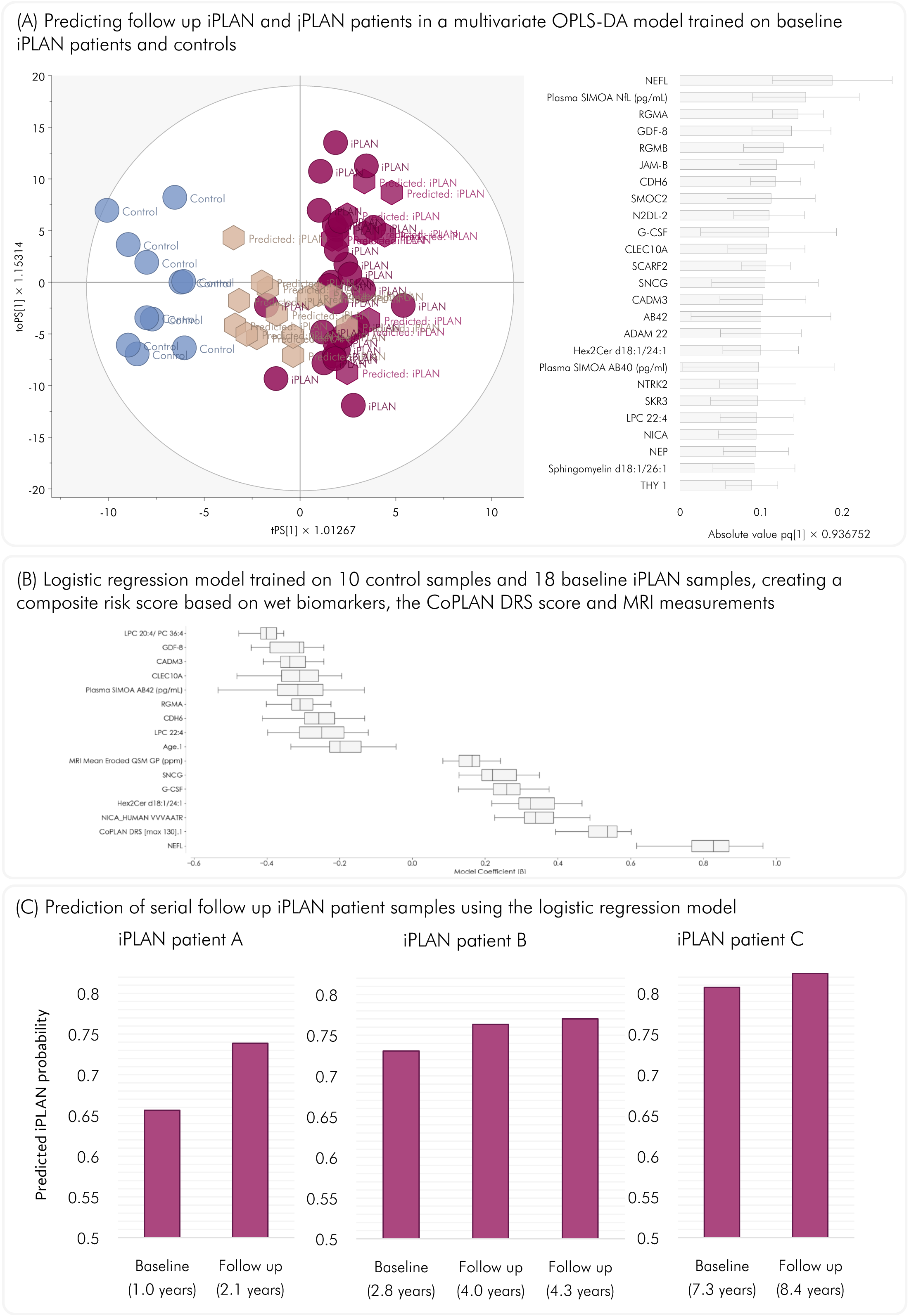
Utilising machine learning to identify the best biomarkers, develop a predictive model and build a portfolio of quantitative metrics for PLAN. **(A)** Predicting follow-up iPLAN and jPLAN patients in a multivariate orthogonal projection to latent structures discriminant analysis (OPLS-DA) analysis trained on baseline iPLAN versus controls (p = 2.9 E^-8^) for proteomic and lipidomic analysis. <u>Left graph:</u> Score scatter plot showing the discrimination between the modelled groups iPLAN (maroon circled) and control (blue circles) as well as the predicted iPLAN follow-up samples (maroon hexagons) and jPLAN (brown hexagons) samples. The horizontal direction of the scatter plots captured the variations **between** PLAN and controls and the vertical dimension capture variations **within** each group. The 11 follow up iPLAN samples projected in the model were all clearly classified as iPLAN. Prediction of samples from the jPLAN group resulted in five out of 14 samples classified as iPLAN. The remaining samples were unclassified by the model, thus not falling firmly into either the control or iPLAN space. <u>Right graph:</u> The protein and lipid biomarkers that best discriminate between iPLAN and controls, ranked in order of significance. **(B)** A logistic regression model trained to discriminate between iPLAN and control samples. The model was based on the CoPLAN-DRS score, the LPC 20:4/PC 36:4 ratio, Quantitative Susceptibility Mapping (QSM) values of the globus pallidus as measured on MRI and the most discriminating wet biomarkers from the OPLS-DA model which remained after we excluded collinear variables (Spearman rho < 0.9 set as limit). Cross validation demonstrated perfect classification metrics with precision, recall, F1 score, and Matthew’s correlation coefficient all 1 ± 0. **(C)** Prediction of longitudinal follow up using the logistic regression model. Longitudinal samples from three individuals excluded from the model training were used for prediction. These consisted of two individuals with two timepoints (patients A and C), and one with three timepoints (patient B). The prediction of these individuals demonstrated a lower prediction score for the baseline sample compared to the follow up samples in all three individuals.

A logistic regression model was developed using the most discriminative, non-collinear wet biomarker variables from the OPLS-DA analysis. Model output probabilities were post-hoc scaled using a continuous age-based multiplier, which gently attenuated scores in younger individuals and reached a plateau of full weighting by age 18. Trained on 28 baseline samples and tested on follow-up iPLAN samples, the model classified all test samples as iPLAN with high confidence (mean probability 0.98 ± 0.020, age-scaled 0.77 ± 0.039). Among jPLAN samples (n = 12), all were classified as iPLAN, though with lower average probability (0.86 ± 0.10, age-scaled 0.72 ± 0.098) and greater variance (P=0.00040). To enhance biological interpretability, CoPLAN-DRS score, MRI QSM GP, and the lyso-PC 20:4/PC 36:4 ratio were incorporated, yielding comparable classification performance. Stratified cross-validation (n = 40; 3 splits, 10 repetitions) demonstrated robust metrics: precision 0.99 ± 0·032, recall 0.98 ± 0.036, F1 score 0.98 ± 0.036 and Matthew’s correlation coefficient 0.96 ± 0.085. A reduced model comprising 15 variables and age achieved perfect classification (all metrics 1 ± 0), with no difference in predicted scores between full and reduced models (**Figure 3B**). In three longitudinal cases, follow-up samples consistently yielded higher probability estimates than baseline, mirroring CoPLAN-DRS score progression (**Figure 3C**). By integrating logistic regression outputs with CoPLAN-DRS score, biomarkers, and QSM, the Digital Disease Dashboard (DDD) was developed to support clinical interpretability and visualise individual progressive disease trajectories over time (**Figure 4**).

**Figure 4:**
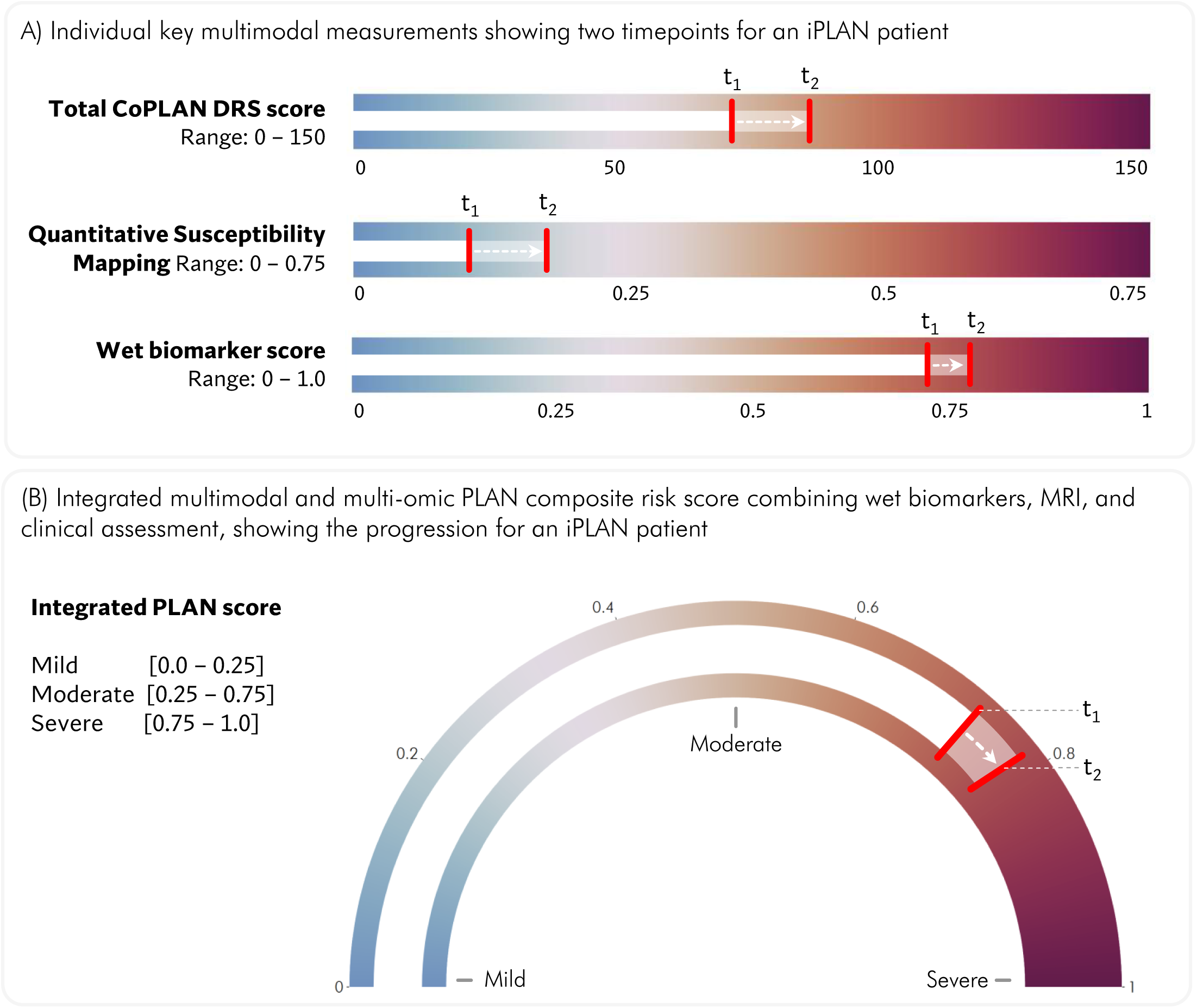
A Digital Disease Dashboard (DDD) allows for objective measurements and quantitative monitoring of disease progression over time. **(A)** The panel shows three informative individual multimodal measurements at 2 timepoints:

- Clinical: Total CoPLAN-DRS score
- Neuroradiological: Quantitative Susceptibility Mapping (QSM) values of the globus pallidus (GP) as measured on MRI (ppm)
- Wet biomarker score, consisting of the 20 most influential proteins and lipids measured in plasma and dried blood spots Visualisation of measurements at baseline time point 1 (t_1_) and subsequent interval measurement at time point 2 (t_2_) allows for monitoring of quantitative metrics longitudinally over time in individual patients. **(B)** The panel shows the composite risk score based on our logistic regression model which uses a combination of 15 intelligently selected variables, combining wet biomarkers, clinical assessment (CoPLAN-DRS score) and neuroradiology (QSM MRI). A higher score indicates worsening disease severity and progression. Visualisation of measurements at baseline time point 1 (t_1_) and subsequent interval measurement at time point 2 (t_2_) allows for monitoring of quantitative metrics longitudinally over time in individual patients.

## DISCUSSION

The PLAN-NHiS represents the largest international PLAN cohort to date, providing key data on motor regression and survival, which will be invaluable for future clinical trials. Historically, two other smaller scale natural history studies that only included infantile PLAN patients have been undertaken, corroborating findings in our study: firstly, a prospective, observational longitudinal study (n = 40) which enrolled patients over a 5 year period^13^; and secondly, a study (n = 28) which included retrospective review of medical records combined with examinations performed at baseline and then follow-up visits at unspecified intervals^14^. While both studies have made important contributions to the field, limitations include small patient cohort size, number of participant centres, and potential caregiver recall bias. Although such challenges are commonly encountered in rare disease natural history studies, our PLAN-NHiS has included a significantly larger number patients with wider geographical reach, representing all PLAN phenotypes, with clinician-reported datasets, in order to provide a broader cross-section of objective patient data from all stages of disease progression. Future large-scale prospective natural history studies in PLAN will further contribute to understanding the natural history of this disease.

Given that existing scales do not fully capture the key disease features of PLAN, we believe that first-in-human clinical trials will benefit from the new CoPLAN-DRS, with its high inter-rater reliability, holistic assessment of multisystemic disease features, and longitudinal monitoring of disease progression. By co-developing the scale with clinical experts and PLAN families, it captures clinically relevant, meaningful measures of disease severity. We identified a strong positive correlation between CoPLAN-DRS scores and age, a surrogate for disease progression given the neurodegenerative nature of PLAN. Through longitudinal assessments, we can track an individual patient’s PLAN disease course over time and quantitatively differentiate disease progression between iPLAN and clinically milder jPLAN patients. Importantly, there are clear examples where a disease-specific rating scale has been instrumental in facilitating trials and regulatory approvals for disease-modifying treatments, including the Children’s Hospital of Philadelphia Infant Test of Neuromuscular Disorders (CHOP INTEND) for RNA therapies for spinal muscular atrophy^15, 16^ and Unified Batten Disease Rating Scale (UBDRS) for enzyme replacement therapy in neuronal ceroid lipofuscinosis^17^.

Basal ganglia iron accumulation is a hallmark of all forms of PLAN and cerebellar atrophy is a near universal finding in iPLAN.^4,18^ Nevertheless, there have been very limited studies to date where such radiological features have been formally quantified, for both PLAN^19^ and other NBIA disorders^20^. We identified a strong positive correlation between QSM values and age, suggesting incremental iron accumulation over time in PLAN. The cerebral and cerebellar volumes were also significantly reduced compared to controls. The longitudinal nature of our study allowed us to identify an incremental increase in iron and reduction in cerebellar volume over time, both on an individual patient and PLAN population basis.

Like many rare genetic disorders, prior to this study, there were no readily available biomarkers for PLAN to aid diagnosis and accurately reflect disease stage. By using DBS for biomarker discovery, we identified proteins and lipids that would have been missed on traditional cell-free plasma or serum. Capturing the red blood cell and leukocyte fraction in DBS allowed us to study intracellular omics, membrane lipids, and the immune system’s response to disease.

Using unbiased mass spectrometry-based proteomics, we found that nicastrin, one of four components of the γ-secretase complex^21^, is upregulated in PLAN. To our knowledge, nicastrin has not been readily detected in human studies. The γ-secretase complex is involved in release of amyloid beta (Aβ) protein and amyloid plaques, a central tenet of Alzheimer’s disease (AD) pathogenesis.^21^ Nicastrin hinders γ-secretase complex substrate interaction^22^, aligning with our findings of reduced Aβ40 and Aβ42 levels in PLAN. The upregulation of nicastrin in PLAN uncovers a new and potentially important disease mechanism beyond tau pathology.^8^ This is particularly noteworthy, given increasing evidence that in AD, tau and Aβ have a synergistic interaction.^23^ Nicastrin may therefore potentially be a biomarker for AD, other tauopathies and non-genetic adult-onset neurodegenerative disorders. Importantly, nicastrin and other protein/lipid biomarkers were validated through DBS mass spectroscopy, which only requires a single drop of blood, easing the burden of sample collection, storage, and transportation – all key advantages for rare paediatric disorders like PLAN.

*PLA2G6* mutations causes phospholipase-A2 enzyme deficiency, impairing conversion of phosphatidylcholine to lysophosphatidylcholine. We developed an assay to measure both substrate and product of this pathway. PLA2G6 deficiency should reduce circulating lysophosphatidylcholine, making it a strong candidate biomarker. We also established a comprehensive lipidomic panel encompassing phospholipids, plasmalogens, and glycolipids, to assess potential downstream or off-target effects of lipid dysregulation. This identified several altered lipids, most notably a strong difference between iPLAN patients and controls in their ratio between the lysolipid lyso-PC C20:4 and its theoretical diacylated precursor PC C36:4. We also found lower levels of lignoceric acid glycosphingolipids and elevated levels of nervonic acid sphingolipids iPLAN versus controls. These findings suggest that PLAN impacts multiple lipid pathways as well as the main PLA2G6 substrate phosphatidylcholine, perhaps in a side chain-specific manner, delineating a broader lipidomic disturbance in PLA2G6 deficiency and underscoring the value of multiplexed lipid profiling for clinical stratification.

Finally, we tested for established biomarkers validated in adult neurodegeneration^24, 25^ using ultra-sensitive Simoa^®^ and Olink^®^ platforms. NfL, a neuronal-specific biomarker that rises with neuroaxonal damage^26^, was significantly raised in PLAN CSF and plasma compared to controls. Elevated NfL levels has been found in other neurodegenerative diseases (including Alzheimer’s disease^27^ and Huntington’s disease^28^), tending to be either stably elevated or increasing as the disease progresses.^26–28^ Intriguingly in PLAN, plasma NfL levels *decrease* over time, though consistently remain higher than controls. It may be postulated that an acute neurodegenerative or neuroinflammatory process earlier in disease leads to initially very high levels of NfL, which then decreases over time, possibly because progressive neurodegeneration leaves proportionately fewer neurons expressing NfL towards end stage disease.

Our proof-of-principle MMM pipeline was built using a well-characterised international cohort with strong patient and public involvement and cross-disciplinary research. This foundation enabled extraction of maximum data from each patient and the most informative features from hundreds of variables, yielding a compact set of optimal biomarkers that reliably predict disease phenotype and progression. The MMM approach objectively identified the best markers of PLAN disease progression: (i) Clinical outcome measures: CoPLAN-DRS score, survival, time to loss of ambulation (ii) Neuroimaging features: QSM MRI values in GP (iii) Biomarkers: NfL and nicastrin. Importantly, these markers have potential clinical utility as efficacy outcome measures for future clinical trials. Furthermore, our MMM approach integrates multimodal data, quantifying disease severity and progression.

By combining clinical measures, neuroimaging signatures, and blood biomarkers into a single machine learning model, we show how complex data can be distilled into a powerful diagnostic and prognostic tool. Using the top discriminatory biomarkers from our multiomics analysis, we developed a machine learning tool that can now (i) distinguish PLAN patients from healthy controls, which could potentially aid clinical diagnosis when genetic testing identifies *PLA2G6* variants of uncertain significance (ii) provide an objective score to accurately classify patients into phenotypic subgroups for prognostication and (iii) track an individual’s disease over time, aiding clinicians/researchers in evaluating the efficacy of therapeutic interventions. Age-based scaling was introduced post-hoc to reflect the lower clinical relevance of biomarker positivity in younger individuals, ensuring that predicted probabilities more accurately align with age-stratified disease risk and reducing the likelihood of overestimation in low-prevalence cohorts. In this era of precision medicine, the MMM approach improves sensitivity and specificity, enables patient stratification and supports longitudinal assessment of treatment response. To translate these findings into a bedside tool, we developed the DDD, an intuitive graphical platform, which condenses multi-dimensional data into a single, interpretable index of patient status. Much like a car dashboard integrates signals from multiple sensors into simple dials like the speedometer, the DDD condenses complex biological data into a clear visual guide, enabling real-time interpretation and decision making.

In conclusion, our MMM approach identifies robust clinical, radiological, and biochemical markers that distinguish PLAN phenotypes and capture the dynamic trajectory of disease progression over time. This strategy can generate big datasets from small cohorts and, through machine learning, enable objective, data-driven insights, even in rare disorders. By integrating diverse data streams into the DDD, a single visual tool, this approach represents more than incremental progress where conventional biomarkers fall short; it unlocks synergistic combinations that transcend the limits of any single modality, offering a paradigm shift in how complex diseases are detected, understood, and managed. The DDD bridges complex bioinformatics with bedside care, while the broader MMM framework holds promise for rare and common diseases alike – accelerating clinical trial readiness, improving patient stratification, and advancing precision-guided therapies.

## Data Availability

All data produced in the present study are available upon reasonable request to the authors.

## ACKNOWLEDGEMENTS

The authors would like to thank all the PLAN patients, families, clinicians and research collaboraters for their invaluable contributions to this study. This study was funded by the National Institute for Health and Care Research (NIHR) Great Ormond Street Hospital Biomedical Research Centre (GOSH BRC) and Cure INAD UK Charity.

